# Visual acuity after proton beam therapy for uveal melanoma: development and validation of a prediction model

**DOI:** 10.64898/2026.10.28.26366694

**Authors:** L.J. Pors, M. Marinkovic, T.H.K. Vu, D. Mohabati, J.C. Bleeker, G.P.M. Luyten, A. Schalenbourg, A. Pica, J. Hrbacek, J.W.M. Beenakker, C.R.N. Rasch, N. Horeweg

## Abstract

**Aim:** To develop proton beam therapy treatment planning dose constraints for uveal melanoma (UM), more knowledge is needed on the impact of dose on toxicity. Here, we identify risk factors and develop prediction models for lower post-therapeutic visual acuity.

**Methods:** LUMC patients treated at Holland PTC between 2020-2023 (n=135) and at PSI between 1987-2019 (n=100) for UM were included. Candidate predictors for severe and moderate to severe vision impairment (Snellen <0.1 and <0.3) were established through literature review and expert interviews and selected through univariable analysis. Multivariable Cox models were fitted on a randomly selected 2/3^rd^ of the combined dataset (n=156). Performance of both models was calculated on the training set and on the held-out validation set (n=79).

**Results:** The final models included baseline visual acuity, ciliary body dose and tumour location for both visual acuity cut-offs, supplemented by the optic disk dose and T-stage for visual acuity <0.1 and macula dose for visual acuity <0.3. Internal performance was good (c-indices 0.760 and 0.761 for visual acuity <0.1 and <0.3, respectively). In the held-out validation set, performance was acceptable (c-indices 0.657 and 0.689) and calibration was insufficient for the model predicting visual acuity <0.1, but acceptable for the model predicting visual acuity <0.3.

**Conclusion:** We developed a model predicting moderate to severe visual impairment with good internal performance and acceptable discrimination and calibration on a held-out test set. This is an important step towards NTCP modelling and the development of dose constraints for ocular proton therapy.

## Introduction

Proton beam therapy (PBT) is a safe eye-sparing treatment method for large and juxtapapillary uveal melanomas (UM), the most common primary ocular malignancy in adults. The aim of PBT is to achieve local control while sparing the eye and visual acuity^1^. While posttreatment local failure and enucleation risks are low (3-9% and 7-10% after five years, respectively^2^), visual acuity loss is common (5-year risk of visual acuity <0.5 31-84%^3–13^).

In the treatment planning workflow, adequate tumour coverage is first ensured. Then, the eye’s gaze angle is optimized to minimize the dose to a fixed set of organs at risk (such as the optic disk, optic nerve, macula, retina, ciliary body, lens, and cornea)^1^. No formal dose constraints are available for PBT planning for UM. In order to define evidence-based dose constraints, normal tissue complication probability (NTCP) models should be developed^14^ and evaluated in terms of discrimination and calibration. To enable future application of NTCP models, this should be demonstrated in independent, preferably external, validation cohorts, to which the model is naïve^15^. Subsequently, dose constraints should be defined to optimize the treatment and lower the risk of post-therapeutic visual acuity loss^14,16^.

Earlier studies have identified patient characteristics^9,13,17,18^, tumour characteristics^3,5,9,13,17–23^, tumour locations^3,5,8,9,17–19,21,24–27^, tumour toxicities at diagnosis^5,7,9,11,13,17–19,21,24,28^ and doses to ocular organs at risk (OARs)^10,13,17,19,21–25^ that correlated with the risk of post-therapeutic visual acuity loss. However, different methods were used to define visual acuity loss (although all based on the Snellen chart), only seven studies performed multivariable analyses and included OAR doses^10,13,17,18,21–23^. None of these reported internal model performance metrics or performed model validation. Thus, while the outcomes of these studies are insightful, a more extensive evaluation of prediction models is necessary to move towards NTCP modelling and subsequently towards evidence-based dose constraints for treatment planning.

Therefore, in this study, we build and validate multivariable prediction models for two different cut-offs for visual acuity after PBT for UM using two combined retrospective, previously published datasets^10,29^. We report the final models, their internal performance metrics calculated on the training dataset, and their performance in a held-out validation dataset.

## Methods

### Patient cohorts

Two previously described cohorts of UM patients of Leiden University Medical Centre (LUMC) were included for analysis. LUMC is a Dutch tertiary referral centre, treating approximately 90% of uveal melanomas in the Netherland. LUMC currently offers Ruthenium-106 brachytherapy (∼50% of the patients), proton beam therapy (∼25%), or enucleation (∼25%). The first cohort initially contained all 103 consecutive LUMC UM patients treated between 01-1987 and 12-2019 at the Paul Scherrer Institute (PSI) in Villigen, Switzerland^10,29^. The second cohort contained all 135 consecutive LUMC UM patients treated between 01-2020 and 12-2023 with PBT at the Holland PTC in the Netherlands^29^. Ethical approval was obtained from our local IRB board (nWMO-D4-2025-009).

### Diagnosis, treatment and follow-up

The UM diagnosis was made based on ultrasound imaging, fundoscopy, fluorescence angiography, and MRI (when available). Patients were eligible for PBT if the tumour prominence (including the sclera) or diameter exceeded 7 or 16 mm, respectively, or if the tumour was juxtapapillary (within one disk diameter from the optic disk). Before 2020, PBT was offered to eligible patients at PSI in Switzerland. However, due to the travel burden, some PBT-eligible patients received Ruthenium-106 brachytherapy or enucleation instead, after shared decision-making^30^. Patients were included for this study if they were ≥18 years old, had no metastases at the time of presentation, and had received no previous primary treatment for UM. Typically, four tantalum markers were placed for tumour definition and position verification. In the first cohort, the marker placement was performed at the Jules Gonin Eye Hospital in Lausanne, Switzerland, and the irradiation at PSI. In the second cohort, the marker placement was performed at LUMC and irradiation at the Holland PTC. 60 CGE irradiation was delivered in four fractions in all patients. A uniform margin of 2.5 mm to the target volume was applied. The ophthalmological follow-up consisted of four visits to our outpatient clinic in the first year, two in the second year, and yearly thereafter, until five years posttreatment. Prophylactic bevacizumab injections were administered from 2012 onwards29.

### Data collection

Data was collected retrospectively from the electronic patient records into pre-defined electronic case report forms in Castor EDC. The tumour size was based on the ultrasound, or, if the ultrasound was not reliable, on the MRI (when available). Tumour prominence was measured including the sclera. Best-corrected visual acuity was measured using the Snellen chart. For the dose parameters, only the V20 (volume of the OAR receiving 12 Gy – 20% of the total dose), V50 and V90 for the various OARs were available from both planning systems (Eyeplan for the first cohort, Eclipse Ocular Proton Planning for the second cohort).

### Definitions and data preparation

To define the most relevant endpoints for visual acuity, and to capture all risk factors, we performed a review of the literature and conducted interviews with six expert clinicians (5 ocular oncologists and 1 radiation oncologist) at our centre. Based on these, we decided to build 2 prediction models for visual acuity, adopting the ICD-11 classification for lasting severe or moderate to severe vision impairment (Snellen below 0.1 and 0.3 still present at the end of follow-up), registered in the first five years after therapy. After eye enucleation, the visual acuity was imputed at 0. Subsequently, the follow-up was truncated at three years posttreatment. Multiple patient, tumour, eye and dose characteristics were identified as candidate predictors (supplementary table 2).

Before the data analysis, the TNM stage was grouped into four categories and into a binary variable (T1-2 vs T3-4). Tumour location was recorded as central, midperipheral, or peripheral using a previously described method^10,29,30^, and subsequently binned in multiple ways, after the expert interviews. From the first cohort of 104 eyes (103 patients), one patient was excluded because they had received PBT for a tumour recurrence, one had objected against further use of their data, and two were excluded for lack of data, leaving 99 patients and 100 treated eyes in this dataset. In the second cohort, there were no missing data for the candidate predictors and all patients remained in the analysis. The two cohorts, without any missing data, were combined and were thereafter randomly split into two groups, stratified by treating centre: a training set and a held-out validation set, using a 2:1 ratio to maintain adequate amounts of events for fitting and validation in the respective sets.

### Statistical analyses

The objective of this study was to build models that could predict visual outcomes, not to assess the aetiology of visual acuity loss after therapy. To pre-select predictors for the models for visual acuities of <0.1 and <0.3, univariable Cox regression models were fitted for all candidate predictors, with a p-value <0.1 as a selection threshold. If multiple dose statistics for one OAR had p<0.1, the dose parameter with the strongest correlation with the visual outcome was selected. Subsequently, Spearman correlation coefficients were calculated for the selected predictors to assess multicollinearity, using a threshold of <0.7. This way, sets of candidate predictors were defined for entry into multivariable regression. Then, backwards stepwise multivariable regression analysis was performed with these sets of candidate predictors, retaining all variables with p-values <0.1. Lastly, the best model for each visual acuity endpoint was chosen based on the -2 log-likelihood of the resulting models.

For each of the final models for the two endpoints, the internal performance was assessed using the c-index, integrated Brier score, and reference Brier score (the latter based on the Kaplan-Meier analysis). Subsequently, the two models (for visual acuity below 0.1 and 0.3) were applied to the held-out validation set. Model performance was calculated using the c-index and the integrated Brier score, and calibration graphs were plotted. The c-index on the held-out test set is considered the primary outcome measure.

To assess differences between the training and test sets, the Chi square test was used for categorical variables, and the Mann-Whitney-U test was used for continuous variables; p-values <0.05 were considered statistically significant. Follow-up times were calculated using the reverse Kaplan-Meier method. Data pre-processing was performed in Python 3.11. Cox regression models were built in SPSS version 29, and validated in R 4.5.1, using the RiskRegression and Survival packages.

## Results

### Cohort description

156 patients were assigned to the training set and 79 patients in the validation set (table 1 and table 2). The two sets were similar, although the macula doses were slightly higher in the training set. In the full cohort, the median age was 62 years, median baseline visual acuity was 0.7, and the median tumour prominence and diameter were 7.9 mm and 16.5 mm, respectively (range 1.5-17.7 and 5.5-25.7, respectively). 79 (34%) tumours were centrally, and 81 (34%) juxtapapillary located. Compared with patients treated at HollandPTC, those treated at PSI were somewhat younger, had slightly larger tumours and somewhat higher doses to the retina and ciliary body but lower lens doses (supplementary table 1).

**Table 1.** Characteristics of the training and validation cohorts.

|  | **All patients**<br>**n=235 (100%)** | **Training cohort**<br>**n=156 (66%)** | **Validation cohort**<br>**n=79 (34%)** | **p-value \*** |
| --- | --- | --- | --- | --- |
| **Age** | 62 (23-88) | 62 (23-84) | 61 (25-88) | 0.35 |
| **Sex - Male** | 122 (52%) | 81 (52%) | 41 (52%) | 1.00 |
| **Diabetes** | 27 (11%) | 15 (10%) | 12 (15%) | 0.21 |
| **Affected eye - right** | 127 (54%) | 78 (50%) | 49 (62%) | 0.08 |
| **Baseline visual acuity - Snellen** | 0.7 (0.0-1.5) | 0.7 (0.0-1.5) | 0.8 (0.0-1.5) | 0.94 |
| **Tumour prominence, including sclera, in mm** | 7.9 (1.5-17.7) | 7.9 (1.7-15.9) | 8.0 (1.5-17.7) | 0.98 |
| **Tumour diameter, in mm** | 16.5 (5.5-25.7) | 16.3 (5.5-25.7) | 16.6 (6.4-24.8) | 1.00 |
| **T stage - AJCC version 8** |  |  |  | 0.34 |
| T1 | 11 (5%) | 6 (4%) | 5 (6%) |  |
| T2 | 56 (24%) | 37 (24%) | 19 (24%) |  |
| T3 | 115 (49%) | 82 (53%) | 33 (42%) |  |
| T4 | 53 (23%) | 31 (20%) | 22 (28%) |  |
| **Location tumour~** |  |  |  | 0.37 |
| Central | 79 (34%) | 57 (37%) | 22 (28%) |  |
| Midperipheral | 123 (52%) | 77 (49%) | 46 (58%) |  |
| Peripheral | 33 (14%) | 22 (14%) | 11 (14%) |  |
| **Juxtapapillary location~** | 81 (34%) | 55 (35%) | 26 (33%) | 0.72 |
| **Central and/or juxtapapillary location** | 172 (73%) | 114 (73%) | 58 (73%) | 0.96 |
| **Treatment site - HollandPTC** | 135 (57%) | 90 (58%) | 45 (57%) | 0.92 |

\* Chi square for categorical variables, Mann-Whitney-U test for continuous variables
~ location of the most central part of the tumour
+ tumour located <1 disk diameter from optic disk
Continuous variables: median (range). In one patient, both eyes were treated. One eye is in the training, and one is in the validation group.

**Table 2.** Proton therapy doses in the training and validation cohorts.

| Covariate | All patients<br>n=235<br>median (range)<br>mean (sd) | Training cohort<br>n=156<br>median (range)<br>mean (sd) | Validation cohort<br>n=79<br>median (range)<br>mean (sd) | p-value* |
| --- | --- | --- | --- | --- |
| **Macula V20 (%)** | 35.7 (0-100)<br>46.3 (46.0) | 50.0 (0-100)<br>50.0 (46.5) | 14.3 (0-100)<br>39.0 (44.3) | 0.07 |
| **Macula V50 (%)** | 0.0 (0-100)<br>39.7 (45.8) | 15.8 (0-100)<br>44.2 (46.8) | 0.0 (0-100)<br>30.9 (42.6) | 0.05 |
| **Macula V90 (%)** | 0.0 (0-100)<br>28.8 (41.8) | 0.0 (0-100)<br>33.4 (43.4) | 0.0 (0-100)<br>19.9 (37.2) | 0.014 |
| **Optic disk V20 (%)** | 33.0 (0-100)<br>49.0 (48.4) | 58.6 (0-100)<br>50.5 (48.0) | 0.0 (0-100)<br>46.1 (49.5) | 0.55 |
| **Optic disk V50 (%)** | 0.0 (0-100)<br>43.9 (47.7) | 14.3 (0-100)<br>44.6 (47.7) | 0.0 (0-100)<br>42.5 (48.1) | 0.62 |
| **Optic disk V90 (%)** | 0.0 (0-100)<br>35.7 (44.6) | 0.0 (0-100)<br>35.7 (44.8) | 0.0 (0-100)<br>35.6 (44.4) | 0.96 |
| **Length of optic nerve**<br>**receiving 20% of dose**<br>**(mm)** | 0.3 (0-10.0)<br>2.1 (2.4) | 0.5 (0.0-10.0)<br>2.2 (2.4) | 0.0 (0-10.0)<br>1.9 (2.4) | 0.29 |
| **Length of optic nerve**<br>**receiving 50% of dose**<br>**(mm)** | 0.0 (0-9.8)<br>1.8 (2.3) | 0.1 (0-9.8)<br>1.9 (2.3) | 0.0 (0-9.8)<br>1.7 (2.2) | 0.47 |
| **Length of optic nerve**<br>**receiving 90% of dose**<br>**(mm)** | 0.0 (0-9.5)<br>1.3 (2.0) | 0.0 (0-9.5)<br>1.3 (2.0) | 0.0 (0-9.3)<br>1.2 (1.9) | 0.91 |
| **Retina V20 (%)** | 37.0 (13.5-72.0)<br>37.1 (9.4) | 37.0 (31.5-63.5)<br>37.3 (8.6) | 36.5 (13.8-72.0)<br>36.7 (10.9) | 0.46 |
| **Retina V50 (%)** | 32.8 (9.7-63.0)<br>33.0 (9.0) | 33.5 (9.7-55.4)<br>33.1 (8.3) | 32.0 (10.3-63.0)<br>32.8 (10.2) | 0.56 |
| **Retina V90 (%)** | 25.5 (3.0-56.0)<br>25.7 (9.8) | 25.3 (5.3-47.1)<br>25.7 (9.0) | 26.0 (3.0-56.0)<br>25.8 (11.3) | 0.93 |
| **Lens V20 (%)** | 44.9 (0-100)<br>45.1 (35.9) | 42.4 (0-100)<br>43.9 (35.1) | 47.0 (0-100)<br>47.4 (37.6) | 0.61 |
| **Lens V50 (%)** | 32.0 (0-100)<br>37.0 (33.8) | 28.3 (0-100)<br>35.4 (33.3) | 38.9 (0-100)<br>40.1 (34.8) | 0.44 |
| **Lens V90 (%)** | 7.4 (0-100)<br>20.8 (26.4) | 4.0 (0-100)<br>18.7 (25.3) | 14.0 (0-100)<br>24.9 (28.1) | 0.09 |
| **Ciliary body V20 (%)** | 38.2 (0.4-99.2)<br>38.0 (17.9) | 37.3 (0.8-92.7)<br>37.9 (16.6) | 40.0 (0.4-99.2)<br>38.4 (20.3) | 0.75 |
| **Ciliary body V50 (%)** | 34.0 (0-96.1)<br>33.6 (16.7) | 33.4 (0-83.0)<br>33.3 (15.5) | 35.0 (0-96.1)<br>34.3 (18.9) | 0.59 |
| **Ciliary body V90 (%)** | 20.7 (0-58.0)<br>19.7 (16.7) | 20.0 (0-56.0)<br>18.4 (16.2) | 24.6 (0-58.0)<br>22.3 (17.4) | 0.18 |

Descriptive statistics of both cohorts. Notably, the macula V20 was 0% in 101 patients and 100% in 87 patients. The macula V90 was 0% in 149 patients and 100% in 45 patients. Similarly, the Optic disk V90 was 0% in 136 patients and 100% in 63 patients.
\* Mann-Whitney-U test

### Visual acuity after therapy

Median follow-up of the combined cohort before truncation was 3.1 years. In both sets, patients experienced a steady, gradual decline of their visual acuity after treatment (figure 1, table 3, supplementary table 2). At three years posttreatment, 85 (54%) patients had severe vision loss (visual acuity <0.1) in the training, compared with 41 (52%) patients in the validation set. Three years after therapy, moderate to severe vision loss (visual acuity <0.3) was found in 105 (67%) patients in the training set and in 50 (63%) in the validation set. After three years, 25 (16%) eyes were enucleated in the training and 12 (15%) in the validation set.

**Figure 1.**
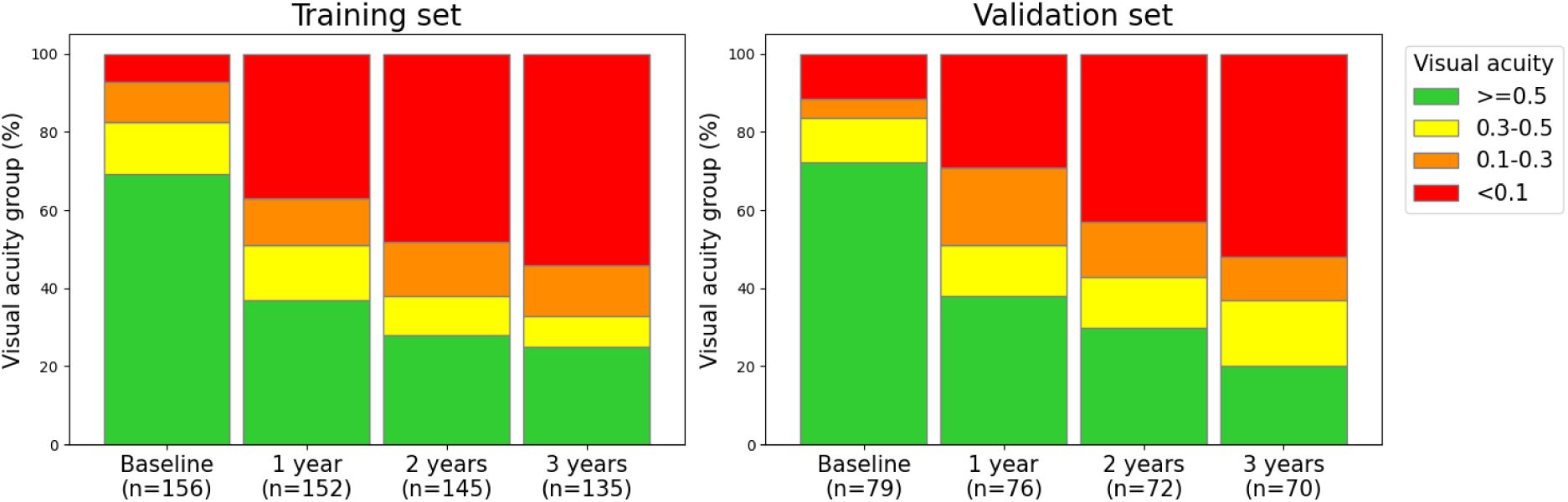
visual acuity in the first three years after therapy in both sets

**Table 3:** Cases with visual acuity below 0.1, 0.3 and 0.5 at 1, 2, and 3 years in the training cohort (n=156)

| **Visual acuity loss cut-off** |  | **First year after therapy** | **First 2 years after therapy** | **First 3 years after therapy** |
| --- | --- | --- | --- | --- |
| **VA loss below 0.1** | *Events (%)* | 57 (37%) | 75 (48%) | 85 (54%) |
|  | *Numbers at risk (%)* | 91 (58%) | 63 (40%) | 33 (21%) |
|  | *Numbers censored (%)* | 8 (5%) | 18 (12%) | 38 (24%) |
| **VA loss below 0.3** | *Events (%)* | 77 (49%) | 97 (62%) | 105 (67%) |
|  | *Numbers at risk (%)* | 75 (48%) | 46 (29%) | 22 (14%) |
|  | *Numbers censored (%)* | 4 (3%) | 13 (8%) | 29 (19%) |
| **Enucleations** | *Numbers enucleated (%)* | 10 (6%) | 20 (13%) | 25 (16%) |

Note – percentages are the % of the original cohort, not the % of the remaining patients

### Prediction model for visual acuity three years after therapy

From the literature search, hypertension, diabetes mellitus, age, tumour size and location, visual acuity, cataract, retinal detachment and glaucoma at baseline, and doses to the macula, optic disk or nerve, and lens were identified as candidate predictors. As outcomes, visual acuity cut-offs below 0.1, 0.3 and 0.5, and the loss of >2 Snellen lines of visual acuity were found. The interviews with the experts established visual acuity cut-offs below 0.1 and 0.3, which are the ICD-11 cut-offs for respectively severe and moderate vision loss^31^, as the most clinically relevant. The experts identified the same candidate predictors as found in the literature and specified the different possible methods to define tumour size and location that were tested (supplementary table 3). As data for pretreatment toxicity was not available for 55 patients treated between 1987-2011, we opted to leave those candidate predictors out of the analyses.

The outcomes of the univariable analyses are shown in supplementary table 3. Baseline visual acuity, tumour location (in multiple definitions), and doses to the macula, optic disk, optic nerve, retina, lens and ciliary body reached the threshold for selection for both models. Additionally, the T-stage and tumour prominence were selected for visual acuity <0.1, and juxtapapillary tumour location was selected for visual acuity <0.3. The correlation coefficients of these variables are shown in supplementary table 4A and 4B. Multivariable Cox’ proportional hazards models were built for all sets of non-colinear variables, which resulted in 8 and 18 models for visual acuity <0.1 and <0.3, respectively.

The final two models with the lowest -2 log-likelihood for each outcome are shown in table 4. The final model for visual acuity <0.1 was constituted of visual acuity at baseline, optic disk V20, ciliary body V20, tumour T-stage, and the location of the most central part of the tumour. For visual acuity <0.3, the model contained the visual acuity at baseline, macula V20, ciliary body V20, and central or juxtapapillary tumour location.

**Table 4:**
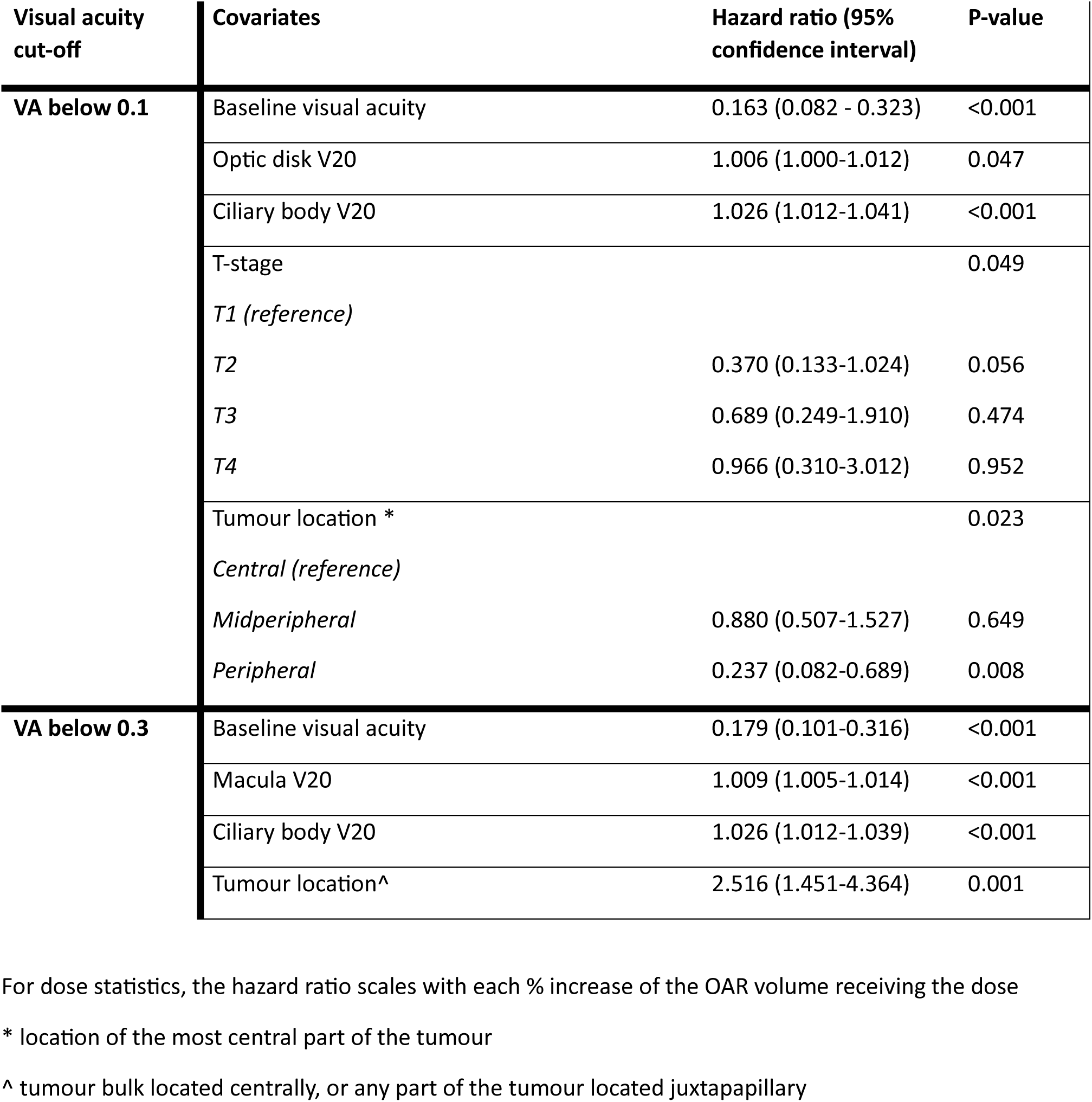
Final prediction models for visual acuity after proton beam therapy built on the training cohort.

### Performance

On the training dataset, the model for visual acuity <0.1 had a c-index of 0.760 (95% confidence interval 0.712-0.808) and an integrated Brier score of 0.161 (reference: 0.227, table 5). The c-index calculated on the validation set was 0.657 (0.561-0.753), and the integrated Brier score was 0.202 (reference: 0.210). The calibration plot for this model shows poor calibration with a systematic underestimation of the risk in the lower risks (0-0.5) and systematic overestimation of the risk in high-risk patients (0.8-1.0) (supplementary figure 1). For the model for visual acuity <0.3, the c-index was 0.761 (0.713-0.809), and the integrated Brier score was 0.152 (reference: 0.228, table 5). On the validation set, the c-index was 0.689 (0.607-0.771), and the integrated Brier score was 0.207 (reference: 0.233). For this model, the calibration shows reasonable calibration, with systematic over- and underestimation of the risk at the ends of the risk spectrum.

**Table 5.** Performance metrics for final prediction models at 3 years of follow-up.

| Visual acuity cutoff | Performance metric | Training set | Validation set |
| --- | --- | --- | --- |
| **VA below 0.1** | C-index (95% confidence interval) | 0.760 (0.712-0.808) | 0.657 (0.561-0.753) |
|  | Integrated Brier score | 0.161 | 0.202 |
|  | Brier score reference* | 0.227 | 0.210 |
| **VA below 0.3** | C-index (95% confidence interval) | 0.761 (0.713-0.809) | 0.689 (0.607-0.771) |
|  | Integrated Brier score | 0.152 | 0.207 |
|  | Brier score reference* | 0.228 | 0.233 |

\* Brier score reference is the Brier score for only the Kaplan Meier analysis

## Discussion

In this study, we found that the visual acuity before treatment, dose to macula, ciliary body and optic disk, and the location and T-stage of the tumour were independent predictors of visual acuity at three years after proton beam therapy. The model for severe vision impairment had a good performance in the training set (c-index 0.76) and an acceptable discrimination (c-index 0.66) but poor calibration in the held-out validation set. The model for moderate to severe vision impairment, however, had a good performance in the training set (c-index 0.76) and acceptable discrimination (c-index 0.69) and calibration in the validation set. This latter model constitutes an important step towards NTCP modelling and the development of dose constraints for ocular proton therapy for uveal melanoma.

Some strengths and weaknesses of this study need to be taken into account when its results are interpreted. The main strengths of this study are the reproducible and structured method for identifying and selecting the candidate predictors and building and validating the prediction models. We used training data from two treatment sites, with sufficient variation in patient selection, beam characteristics, planning software and planning protocols. Additionally, the patients treated at PSI were treated between 1987-2019, leading to some differences in post-treatment follow-up and availability of (prophylactic) therapies for post-treatment toxicities, such as intravitreal injections^32^. The heterogeneity of the training data may add to the applicability of the prediction models elsewhere. Additionally, the organ at risk doses are based on universal geometrical eye models adapted to the patient but not based on ocular imaging and are therefore bound to some uncertainty. However, as most patients treated with ocular proton therapy are treated using geometrical models^33^, the use of such model-based organ at risk doses adds to the applicability of the prediction model elsewhere. The retrospective data collection may have led to underestimation of some data, such as comorbidities at baseline and post therapeutic vision loss, due to underreporting. Finally, the relatively small size of the training set might have led to missing true effects, which we aimed to counter with a threshold for variable selection of p<0.1.

As in our study, previous studies found that tumour size^7,9,11,18,21–23^ and location^9,11,17,18,21^, visual acuity before treatment^7,11,13,18,21,23^, and the dose to the macula^10,13,21–23^ and optic disk^10,17,21,22^ or nerve^10,13^ were independent predictors of visual acuity after proton therapy in multivariable models. Notably, the strongest predictor in both prediction models in this study was the pre-treatment visual acuity. This should be considered during patient counselling. Unlike in our study, previous studies also identified age^9,13,17,23^ and diabetes^7,11,13,18,21,23^ as predictors of posttreatment visual acuity. In our study, these were not predictive of loss of visual acuity <0.1 and <0.3. We identified a correlation between ciliary body dose and loss of visual acuity that has not been described earlier. The ciliary body dose had a strong correlation with the dose to the lens (Spearman’s ρ >0.8), which has previously been described to impact visual acuity in a study where ciliary body dose was not evaluated^21^, but in our analysis ciliary body dose was a stronger predictor than lens dose. The ciliary body dose may be higher in some patients where a lower visual acuity can be expected: those with a high dose to either the lens or the complete eye, or those where in treatment planning, a large volume of the ciliary body receiving a high dose is accepted to minimize posterior pole dose in tumours close to the macula or optic disk. Further research into this relation is warranted, as the dose effects observed in this study may partly reflect tumour position or planning trade-offs.

Currently, no specific dose constraints exist for organs at risk in uveal melanoma proton therapy planning. If prediction models are used in clinical practise, they should be able to accurately predict the risk of the clinical outcome, and therefore, the accuracy of their predictions should be assessed^34^. Earlier studies have used multivariable analyses to identify predictors of vision loss after ocular proton therapy^10,13,17,18,21–23^. One study by Espensen et al. took prediction modelling one step further by using LASSO for variable selection and presented two NTCP models: a model for >0.3 LogMAR visual acuity deterioration from baseline and a model for loss of good pretreatment visual acuity. Unfortunately, internal performance was not reported and discrimination and calibration metrics on a held-out or external set were not calculated^22^. In our study, we build on this by presenting two prediction models and reporting their respective performance in the training and in a held-out validation set. The model for severe vision impairment had a good performance in the training set and an acceptable discrimination but poor calibration in the held-out validation set. This model should therefore not be used for NTCP modelling. The model for moderate to severe vision impairment, however, had a good performance in the training set and acceptable discrimination and calibration on the validation set. To facilitate external validation of this model, we published the model’s coefficients, covariance matrix and the baseline hazards in supplementary tables 5, 6 and 7. If the external performance of the model is acceptable, NTCP models can be presented based on these prediction models, and dose constraints for ocular proton therapy treatment planning can be developed.

Future studies could also consider developing new prediction models with better performance. In any case, we recommend using a transparent and systematic workflow, like in our study. Five possible strategies aiming at a better model performance could be considered. First, new models should be built based on larger, international cohorts, with sufficient variation regarding patient, tumour and treatment characteristics. Second, additional data could be included in these models, such as pre-treatment toxicity, full dose volume histogram curves of the doses to the organs at risk, and imaging data (such as OCT images, MR-images or fundus photographs). Third, statistical methods such as bootstrapping, shrinkage or LASSO could be implemented. Fourth, organ at risk doses could be based on imaging-based eye models (ideally including MRI, OCT, and fundus imaging), to improve the accuracy of the organ at risk locations and thus the estimated doses to these structures. Finally, some candidate predictors may have better predictive performance after transformation (e.g. by using LogMAR for visual acuity at baseline).

In conclusion, we have developed two prediction models for loss of visual acuity after proton beam therapy for uveal melanoma. One of these models, predicting moderate to severe visual impairment, showed good internal performance and acceptable discrimination and calibration on a held-out test set. The predictors in the model are known to be associated with loss of visual acuity: the visual acuity before treatment, dose to macula, ciliary body and tumour location. This is an important step towards NTCP modelling and the development of dose constraints for ocular proton therapy.

## Data Availability

Relevant data is available in the manuscript and supplementary material.

## Notes

### Competing Interest Statement

Funding for this project has been provided by the Dutch Cancer Society (KWF)

### Author Declarations

IRB of the Leiden University Medical Centre gave ethical approval for this work (nWMO‐D4‐2025‐009)

### Funding

- Dutch Cancer Society: KWF 13400

## References

1. Hrbacek J, Kacperek A, Beenakker J-WM, et al. PTCOG Ocular Statement: Expert Summary of Current Practices and Future Developments in Ocular Proton Therapy. International Journal of Radiation Oncology*Biology*Physics. 2024;doi:10.1016/j.ijrobp.2024.06.017

2. Verma V, Mehta MP. Clinical Outcomes of Proton Radiotherapy for Uveal Melanoma. Clin Oncol (R Coll Radiol). Aug 2016;28(8):e17–27. doi:10.1016/j.clon.2016.01.034

3. Gragoudas ES, Seddon JM, Egan K, et al. Long-term results of proton beam irradiated uveal melanomas. Ophthalmology. Apr 1987;94(4):349–53. doi:10.1016/s0161-6420(87)33456-6

4. Gragoudas ES, Lane AM, Munzenrider J, Egan KM, Li W. Long-term risk of local failure after proton therapy for choroidal/ciliary body melanoma. Trans Am Ophthalmol Soc. 2002;100:43–8; discussion 48-9.

5. Damato B, Kacperek A, Chopra M, Campbell IR, Errington RD. Proton beam radiotherapy of choroidal melanoma: the Liverpool-Clatterbridge experience. Int J Radiat Oncol Biol Phys. Aug 1 2005;62(5):1405–11. doi:10.1016/j.ijrobp.2005.01.016

6. Mosci C, Lanza FB, Barla A, et al. Comparison of clinical outcomes for patients with large choroidal melanoma after primary treatment with enucleation or proton beam radiotherapy. Ophthalmologica. 2012;227(4):190–6. doi:10.1159/000334401

7. Patel AV, Lane AM, Morrison MA, et al. Visual Outcomes after Proton Beam Irradiation for Choroidal Melanomas Involving the Fovea. Ophthalmology. Feb 2016;123(2):369–377. doi:10.1016/j.ophtha.2015.09.031

8. Papakostas TD, Lane AM, Morrison M, Gragoudas ES, Kim IK. Long-term Outcomes After Proton Beam Irradiation in Patients With Large Choroidal Melanomas. JAMA Ophthalmol. Nov 1 2017;135(11):1191–1196. doi:10.1001/jamaophthalmol.2017.3805

9. Toutee A, Angi M, Dureau S, et al. Long-Term Visual Outcomes for Small Uveal Melanoma Staged T1 Treated by Proton Beam Radiotherapy. Cancers (Basel). Jul 24 2019;11(8)doi:10.3390/cancers11081047

10. Marinkovic M, Pors LJ, Van Den Berg V, et al. Clinical Outcomes after International Referral of Uveal Melanoma Patients for Proton Therapy. Cancers. 2021;13(24):6241. doi:10.3390/cancers13246241

11. Lane AM, Oxenreiter MM, Hashmi M, et al. A Comparison of Treatment Outcomes after Standard Dose (70 Gy) versus Reduced Dose (50 Gy) Proton Radiation in Patients with Uveal Melanoma. Ophthalmol Retina. Nov 2022;6(11):1089–1097. doi:10.1016/j.oret.2022.05.006

12. Egger E, Zografos L, Munkel G, Bohringer T, Bercher L, Chamot L. Results of proton radiotherapy for uveal melanomas. Front Radiat Ther Oncol. 1997;30:111–22. doi:10.1159/000425694

13. Polishchuk AL, Mishra KK, Weinberg V, et al. Temporal Evolution and Dose-Volume Histogram Predictors of Visual Acuity After Proton Beam Radiation Therapy of Uveal Melanoma. Int J Radiat Oncol Biol Phys. Jan 1 2017;97(1):91–97. doi:10.1016/j.ijrobp.2016.09.019

14. Marks LB, Yorke ED, Jackson A, et al. Use of Normal Tissue Complication Probability Models in the Clinic. International Journal of Radiation Oncology*Biology*Physics. 2010;76(3):S10–S19. doi:10.1016/j.ijrobp.2009.07.1754

15. Stieb S, Lee A, van Dijk LV, Frank S, Fuller CD, Blanchard P. NTCP Modeling of Late Effects for Head and Neck Cancer: A Systematic Review. Int J Part Ther. Summer 2021;8(1):95–107. doi:10.14338/20-00092

16. Langendijk JA, Lambin P, De Ruysscher D, Widder J, Bos M, Verheij M. Selection of patients for radiotherapy with protons aiming at reduction of side effects: The model-based approach. Radiotherapy and Oncology. 2013;107(3):267–273. doi:10.1016/j.radonc.2013.05.007

17. Pica A, Weber DC, Vallat L, et al. Good long-term visual outcomes of parapapillary choroidal melanoma patients treated with proton therapy: a comparative study. Int Ophthalmol. Feb 2021;41(2):441–452. doi:10.1007/s10792-020-01594-z

18. Gragoudas E, Li W, Goitein M, Lane AM, Munzenrider JE, Egan KM. Evidence-based estimates of outcome in patients irradiated for intraocular melanoma. Arch Ophthalmol. Dec 2002;120(12):1665–71. doi:10.1001/archopht.120.12.1665

19. Jung SK, Park YH, Shin DH, et al. Visual outcomes of proton beam therapy for choroidal melanoma at a single institute in the Republic of Korea. PLoS One. 2020;15(12):e0242966. doi:10.1371/journal.pone.0242966

20. Weber B, Paton K, Ma R, Pickles T. Outcomes of Proton Beam Radiotherapy for Large Non-Peripapillary Choroidal and Ciliary Body Melanoma at TRIUMF and the BC Cancer Agency. Ocul Oncol Pathol. Sep 2015;2(1):29–35. doi:10.1159/000433546

21. Seddon JM, Gragoudas ES, Polivogianis L, et al. Visual outcome after proton beam irradiation of uveal melanoma. Ophthalmology. May 1986;93(5):666–74. doi:10.1016/s0161-6420(86)33700-x

22. Espensen CA, Kiilgaard JF, Appelt AL, et al. Dose-Response and Normal Tissue Complication Probabilities after Proton Therapy for Choroidal Melanoma. Ophthalmology. Jan 2021;128(1):152–161. doi:10.1016/j.ophtha.2020.06.030

23. Thariat J, Grange JD, Mosci C, et al. Visual Outcomes of Parapapillary Uveal Melanomas Following Proton Beam Therapy. Int J Radiat Oncol Biol Phys. May 1 2016;95(1):328–335. doi:10.1016/j.ijrobp.2015.12.011

24. Fuss M, Loredo LN, Blacharski PA, Grove RI, Slater JD. Proton radiation therapy for medium and large choroidal melanoma: preservation of the eye and its functionality. Int J Radiat Oncol Biol Phys. Mar 15 2001;49(4):1053–9. doi:10.1016/s0360-3016(00)01430-9

25. Riechardt AI, Cordini D, Willerding GD, et al. Proton beam therapy of parapapillary choroidal melanoma. Am J Ophthalmol. Jun 2014;157(6):1258–65. doi:10.1016/j.ajo.2014.02.032

26. Sikuade MJ, Salvi S, Rundle PA, Errington DG, Kacperek A, Rennie IG. Outcomes of treatment with stereotactic radiosurgery or proton beam therapy for choroidal melanoma. Eye (Lond). Sep 2015;29(9):1194–8. doi:10.1038/eye.2015.109

27. Wu F, Lane AM, Trofimov A, Shih HA, Gragoudas ES, Kim IK. Outcomes after Proton Beam Irradiation in Patients with Choroidal Melanoma Eligible for Investigational AU-011 Treatment. Ocular Oncology and Pathology. 2023;9(5-6):152–157. doi:10.1159/000534184

28. Hussain G, Lam J, Eleuteri A, et al. Outcomes Following Treatment with Notched Proton Beams for Peripapillary Choroidal Melanomas. Cancers (Basel). Nov 18 2025;17(22)doi:10.3390/cancers17223684

29. Rasch C, Marinkovic M, Pors L, et al. Three years of ocular proton therapy in the Netherlands, clinical results. Acta Ophthalmol. Jul 2 2026;doi:10.1111/aos.70195

30. Pors LJ, Marinkovic M, Deuzeman HH, et al. Clinical outcomes and risk factors for local failure and visual impairment in patients treated with Ru-106 brachytherapy for uveal melanoma. Clin Transl Radiat Oncol. May 2025;52:100939. doi:10.1016/j.ctro.2025.100939

31. International Classification of Diseases ICD-11 for Mortality and Morbidity Statistics. 9D90 Vision Impairment Including Blindness. Accessed 16-10, 2023. https://icd.who.int/browse11/l-m/en#/http%253a%252f%252fid.who.int%252ficd%252fentity%252f1103667651

32. Mantel I, Schalenbourg A, Bergin C, Petrovic A, Weber DC, Zografos L. Prophylactic use of bevacizumab to avoid anterior segment neovascularization following proton therapy for uveal melanoma. Am J Ophthalmol. Oct 2014;158(4):693–701 e2. doi:10.1016/j.ajo.2014.07.002

33. Mortimer L, Slopsema R, Mazal A, et al. PTCOG Ocular Survey - Perspective on Ocular Particle Therapy: Current Practices and Emerging Trends. Int J Part Ther. Mar 2026;19:101300. doi:10.1016/j.ijpt.2026.101300

34. Riley RD, Archer L, Snell KIE, et al. Evaluation of clinical prediction models (part 2): how to undertake an external validation study. BMJ. 2024;384:e074820. doi:10.1136/bmj-2023-074820

