## Supplementary tables for "Visual acuity after proton beam therapy for uveal melanoma: development and validation of a prediction model"

### Supplementary material

**Supplementary table 1A: comparison between the two cohorts (treated at two different treatment centres)**

| | PSI cohort<br>n=100 | HollandPTC cohort<br>n=135 | p-value * |
|---|---|---|---|
| **Age** | 58 (23-84) | 64 (25-88) | 0.023 |
| **Sex - Male** | 54 (54%) | 68 (50%) | 0.53 |
| **Diabetes** | 8 (8%) | 19 (14%) | 0.15 |
| **Affected eye - right** | 56 (56%) | 71 (53%) | 0.60 |
| **Baseline visual acuity - Snellen** | 0.8 (0.0-1.3) | 0.7 (0.0-1.5) | 0.69 |
| **Tumour prominence, including sclera, in mm** | 8.4 (1.5-17.7) | 7.4 (1.7-15.9) | 0.13 |
| **Tumour diameter, in mm** | 18.7 (6.4-25.7) | 15.0 (5.5-21.6) | <0.001 |
| **T stage - AJCC version 8** | | | <0.001 |
| T1 | 2 (2%) | 9 (7%) | |
| T2 | 26 (26%) | 30 (22%) | |
| T3 | 36 (36%) | 79 (59%) | |
| T4 | 36 (36%) | 17 (13%) | |
| **Location tumour~** | | | 0.46 |
| Central | 30 (30%) | 49 (36%) | |
| Midperipheral | 57 (57%) | 66 (49%) | |
| Peripheral | 13 (13%) | 20 (15%) | |
| **Juxtapapillary location~** | 33 (33%) | 48 (36%) | 0.68 |
| **Central and/or juxtapapillary location** | 75 (75%) | 97 (72%) | 0.59 |

* Chi square for categorical variables, Mann-Whitney-U test for continuous variables

~ location of the most central part of the tumour

+ tumour located <1 disk diameter from optic disk

Continuous variables: median (range). In one patient, both eyes were treated. Therefore, n=99 in the age, sex and diabetes rows for the PSI cohort.

**Supplementary table 1B: proton therapy doses in the two cohorts (treated at two different treatment centres)**

| Covariate | PSI cohort<br>n=100<br>median (range)<br>mean (sd) | HollandPTC cohort<br>n=135<br>median (range)<br>mean (sd) | p-value* |
|---|---|---|---|
| **Macula V20 (%)** | 40.0 (0.0-100.0)<br>46.5 (45.7) | 33.3 (0.0-100.0)<br>46.2 (46.3) | 0.97 |
| **Macula V50 (%)** | 0.0 (0.0-100.0)<br>41.0 (45.5) | 0.0 (0.0-100.0)<br>38.8 (46.1) | 0.79 |
| **Macula V90 (%)** | 0.0 (0.0-100.0)<br>30.8 (41.8) | 0.0 (0.0-100.0)<br>27.4 (41.9) | 0.41 |
| **Optic disk V20 (%)** | 25.0 (0.0-100.0)<br>48.6 (48.8) | 57.1 (0.0-100.0)<br>49.3 (48.4) | 0.97 |
| **Optic disk V50 (%)** | 0.0 (0.0-100.0)<br>44.6 (47.8) | 0.0 (0.0-100.0)<br>43.4 (47.9) | 0.97 |
| **Optic disk V90 (%)** | 0.0 (0.0-100.0)<br>40.5 (47.1) | 0.0 (0.0-100.0)<br>32.1 (42.4) | 0.12 |
| **Length of optic nerve receiving 20% of dose (mm)** | 0.2 (0.0-10.0)<br>2.1 (2.7) | 0.58 (0.0-5.2)<br>2.1 (2.2) | 0.55 |
| **Length of optic nerve receiving 50% of dose (mm)** | 0.0 (0.0-9.8)<br>1.8 (2.5) | 0.0 (0.0-5.2)<br>1.8 (2.1) | 0.71 |
| **Length of optic nerve receiving 90% of dose (mm)** | 0.0 (0.0-9.5)<br>1.5 (2.2) | 0.0 (0.0-5.2)<br>1.1 (1.7) | 0.27 |
| **Retina V20 (%)** | 38.5 (16.0-72.0)<br>38.1 (9.8) | 35.0 (13.5-63.5)<br>36.3 (9.1) | 0.08 |
| **Retina V50 (%)** | 35.0 (14.0-63.0)<br>35.0 (9.2) | 30.6 (9.7-55.4)<br>31.6 (9.2) | 0.002 |
| **Retina V90 (%)** | 31.0 (3.0-56.0)<br>29.8 (10.3) | 22.8 (5.3-47.1)<br>22.7 (8.3) | <0.001 |
| **Lens V20 (%)** | 30.0 (0.0-96.0)<br>34.3 (29.5) | 59.7 (0.0-100.0)<br>53.1 (38.2) | <0.001 |
| **Lens V50 (%)** | 20.5 (0.0-89.0)<br>27.0 (26.6) | 44.4 (0.0-100.0)<br>44.4 (36.6) | <0.001 |
| **Lens V90 (%)** | 7.5 (0.0-79.0)<br>18.3 (22.0) | 7.4 (0.0-100.0)<br>22.6 (29.2) | 0.81 |
| **Ciliary body V20 (%)** | 35.5 (4.0-74.0)<br>35.8 (12.8) | 40.0 (0.4-99.2)<br>39.7 (20.8) | 0.07 |
| **Ciliary body V50 (%)** | 31.0 (3.0-67.0)<br>32.2 (12.9) | 35.0 (0.0-96.1)<br>34.7 (19.0) | 0.24 |
| **Ciliary body V90 (%)** | 25.5 (0.0-58.0)<br>23.5 (16.8) | 18.6 (0.0-52.7)<br>16.9 (16.1) | 0.005 |

* Mann-Whitney-U test

**Supplementary table 2: Cases with visual acuity below 0.1, 0.3 and 0.5 at 1, 2, and 3 years in the test cohort (n=79)**

| Visual acuity loss cut-off | | First year after therapy | First 2 years after therapy | First 3 years after therapy |
|---|---|---|---|---|
| **VA loss below 0.1** | *Events* | 23 (29%) | 34 (43%) | 41 (52%) |
| | *Numbers at risk (%)* | 51 (65%) | 32 (41%) | 20 (25%) |
| | *Numbers censored (%)* | 5 (6%) | 13 (16%) | 18 (23%) |
| **VA loss below 0.3** | *Events (%)* | 39 (49%) | 45 (57%) | 50 (63%) |
| | *Numbers at risk (%)* | 37 (47%) | 25 (32%) | 16 (20%) |
| | *Numbers censored (%)* | 3 (4%) | 9 (11%) | 13 (16%) |
| **Enucleations** | *Numbers enucleated (%)* | 8 (10%) | 12 (15%) | 12 (15%) |

**Supplementary table 3 – Overview of all candidate predictors and selection for prediction modelling based on univariate regression**

| **Candidate predictor** | **Selected for prediction model*** |
|---|---|
| Diabetes mellitus | None |
| Age | None |
| TNM stage (T1 vs T2 vs T3 vs T4) | Visual acuity <0.1 |
| TNM (T1-T2 vs T3-T4) | None |
| Tumour diameter | None |
| Tumour prominence (incl. sclera) | Visual acuity <0.1 |
| Tumour location (bulk, central vs midperipheral vs peripheral) | Visual acuity <0.1, <0.3 |
| Tumour location (bulk, central vs midperipheral or peripheral) | Visual acuity <0.3 |
| Tumour location (most central part, central vs midperipheral vs peripheral) | Visual acuity <0.1, <0.3 |
| Tumour location (most central part, central vs midperipheral or peripheral | Visual acuity <0.1, <0.3 |
| Tumour bulk located midperipheral and superior | None |
| Juxtapapillary location | Visual acuity <0.3 |
| Tumour bulk located centrally or tumour located juxtapapillary | Visual acuity <0.1, <0.3 |
| Macula V20 | Visual acuity <0.1, <0.3 |
| Macula V50 | Visual acuity <0.1, <0.3 |
| Macula V90 | Visual acuity <0.1, <0.3 |
| Optic disk V20 | Visual acuity <0.1, <0.3 |
| Optic disk V50 | Visual acuity <0.1, <0.3 |
| Optic disk V90 | Visual acuity <0.1, <0.3 |
| Optic nerve length 20% | Visual acuity <0.1, <0.3 |
| Optic nerve length 50% | Visual acuity <0.1, <0.3 |
| Optic nerve length 90% | None |
| Retina V20 | Visual acuity <0.1, <0.3 |
| Retina V50 | Visual acuity <0.1, <0.3 |
| Retina V90 | Visual acuity <0.1, <0.3 |
| Lens V20 | Visual acuity <0.1, <0.3 |
| Lens V50 | Visual acuity <0.1, <0.3 |
| Lens V90 | None |
| Ciliary body V20 | Visual acuity <0.1, <0.3 |
| Ciliary body V50 | Visual acuity <0.1, <0.3 |
| Ciliary body V90 | None |
| Baseline visual acuity | Visual acuity <0.1, <0.3 |

* Variables were selected for multivariable analysis if they passed the threshold (p<0.1) in univariable analysis. If multiple dose levels for one OAR passed the threshold, the dose level with the strongest relation to the respective outcome was selected for further modelling (underlined).

**Supplementary table 4A – Correlations for all variables combinations selected for the prediction model for VA <0.1**

| | TNM stage | Tumour location (bulk, 3 categories) | Tumour location (any part, 3 categories) | Tumour location (any part, 2 categories) | Central or juxtapapil-lary tumour location | Tumour prominence | Macula V20 | Optic disk V20 | Optic nerve length 20% | Retina V50 | Lens V50 | Ciliary body V20 | Baseline visual acuity |
|---|---|---|---|---|---|---|---|---|---|---|---|---|---|
| TNM stage | | 0.44 | 0.40 | 0.46 | -0.18 | 0.54 | -0.38 | -0.46 | -0.48 | 0.32 | 0.26 | 0.34 | 0.04 |
| Tumour location (bulk, 3 categories) | 0.44 | | 0.74 | 0.60 | -0.69 | 0.33 | -0.63 | -0.59 | -0.59 | -0.27 | 0.27 | 0.25 | 0.15 |
| Tumour location (any part, 3 categories) | 0.40 | 0.74 | | 0.92 | -0.53 | 0.31 | -0.67 | -0.59 | -0.60 | -0.20 | 0.27 | 0.25 | 0.13 |
| Tumour location (any part, 2 categories) | 0.46 | 0.60 | 0.92 | | -0.31 | 0.34 | -0.62 | -0.54 | -0.56 | -0.11 | 0.19 | 0.20 | 0.18 |
| Central or juxtapapillary tumour location | -0.18 | -0.69 | -0.53 | -0.31 | | -0.20 | 0.48 | 0.49 | 0.48 | 0.27 | -0.31 | -0.29 | 0.00 |
| Tumour prominence | 0.54 | 0.33 | 0.31 | 0.34 | -0.20 | | -0.17 | -0.29 | -0.36 | 0.28 | 0.42 | 0.39 | -0.12 |
| Macula V20 | -0.38 | -0.63 | -0.67 | -0.62 | 0.48 | -0.17 | | 0.67 | 0.66 | 0.41 | -0.14 | -0.18 | -0.16 |
| Optic disk V20 | -0.46 | -0.59 | -0.59 | -0.54 | 0.49 | -0.29 | 0.67 | | 0.94 | 0.34 | -0.22 | -0.25 | -0.13 |
| Optic nerve length 20% | -0.48 | -0.59 | -0.60 | -0.56 | 0.48 | -0.36 | 0.66 | 0.94 | | 0.28 | -0.25 | -0.30 | -0.11 |
| Retina V50 | 0.32 | -0.27 | -0.20 | -0.11 | 0.27 | 0.28 | 0.41 | 0.34 | 0.28 | | 0.06 | 0.18 | -0.12 |
| Lens V50 | 0.26 | 0.27 | 0.27 | 0.19 | -0.31 | 0.42 | -0.14 | -0.22 | -0.25 | 0.06 | | 0.86 | -0.17 |
| Ciliary body V20 | 0.34 | 0.25 | 0.25 | 0.20 | -0.29 | 0.39 | -0.18 | -0.25 | -0.30 | 0.18 | 0.86 | | -0.14 |
| Baseline visual acuity | 0.04 | 0.15 | 0.13 | 0.18 | 0.00 | -0.12 | -0.16 | -0.13 | -0.11 | -0.12 | -0.17 | -0.14 | |

**Supplementary table 4B – Correlations for all variable combinations selected for the prediction model for VA <0.3**

| | Tumour location (bulk, 3 categories) | Tumour location (any part, 3 categories) | Tumour location (bulk, 2 categories) | Tumour location (any part, 2 categories) | Central or juxtapapillary tumour location | Juxtapapillary tumour location | Macula V20 | Optic disk V20 | Optic nerve 20% | Retina V50 | Ciliary body V20 | Lens V50 | Baseline visual acuity |
|---|---|---|---|---|---|---|---|---|---|---|---|---|---|
| Tumour location (bulk, 3 categories) | | 0.74 | 0.71 | 0.60 | -0.69 | -0.50 | -0.63 | -0.59 | -0.59 | -0.27 | 0.25 | 0.27 | 0.15 |
| Tumour location (any part, 3 categories) | 0.74 | | 0.50 | 0.92 | -0.53 | -0.54 | -0.67 | -0.59 | -0.60 | -0.20 | 0.25 | 0.27 | 0.13 |
| Tumour location (bulk, 2 categories) | 0.71 | 0.50 | | 0.55 | -0.05 | -0.37 | -0.39 | -0.37 | -0.41 | -0.02 | 0.06 | 0.05 | 0.22 |
| Tumour location (any part, 2 categories) | 0.60 | 0.92 | 0.55 | | -0.31 | -0.53 | -0.62 | -0.54 | -0.56 | -0.11 | 0.20 | 0.19 | 0.18 |
| Central or juxtapapillary tumour location | -0.69 | -0.53 | -0.05 | -0.31 | | 0.45 | 0.48 | 0.49 | 0.48 | 0.27 | -0.29 | -0.31 | 0.00 |
| Juxtapapillary tumour location | -0.50 | -0.54 | -0.37 | -0.53 | 0.45 | | 0.51 | 0.77 | 0.82 | 0.12 | -0.33 | -0.26 | -0.03 |
| Macula V20 | -0.63 | -0.67 | -0.39 | -0.62 | 0.48 | 0.51 | | 0.67 | 0.66 | 0.41 | -0.18 | -0.14 | -0.16 |
| Optic disk V20 | -0.59 | -0.59 | -0.37 | -0.54 | 0.49 | 0.77 | 0.67 | | 0.94 | 0.34 | -0.25 | -0.22 | -0.13 |
| Optic nerve 20% | -0.59 | -0.60 | -0.41 | -0.56 | 0.48 | 0.82 | 0.66 | 0.94 | | 0.28 | -0.30 | -0.25 | -0.11 |
| Retina V50 | -0.27 | -0.20 | -0.02 | -0.11 | 0.27 | 0.12 | 0.41 | 0.34 | 0.28 | | 0.18 | 0.06 | -0.12 |
| Ciliary body V20 | 0.25 | 0.25 | 0.06 | 0.20 | -0.29 | -0.33 | -0.18 | -0.25 | -0.30 | 0.18 | | 0.86 | -0.14 |
| Lens V50 | 0.27 | 0.27 | 0.05 | 0.19 | -0.31 | -0.26 | -0.14 | -0.22 | -0.25 | 0.06 | 0.86 | | -0.17 |
| Baseline visual acuity | 0.15 | 0.13 | 0.22 | 0.18 | 0.00 | -0.03 | -0.16 | -0.13 | -0.11 | -0.12 | -0.14 | -0.17 | |

**Supplementary table 5: coefficients of both models**

| Visual acuity cut-off | Covariates | Coefficient |
|---|---|---|
| **VA below 0.1** | Baseline visual acuity | -1.81514689 |
| | Optic disk V20 | 0.00613534 |
| | Ciliary body V20 | 0.02576647 |
| | T-stage | |
| | *T1 (reference)* | |
| | *T2* | -0.99545795 |
| | *T3* | -0.37238536 |
| | *T4* | -0.03490722 |
| | Tumour location * | |
| | *Central (reference)* | |
| | *Midperipheral* | -0.12796489 |
| | *Peripheral* | -1.43956509 |
| **VA below 0.3** | Baseline visual acuity | -1.723079925 |
| | Macula V20 | 0.009327241 |
| | Ciliary body V20 | 0.025181967 |
| | Tumour location^ | 0.922830313 |

* location of the most central part of the tumour

^ tumour located centrally and/or juxtapapillary

**Supplementary table 6A: covariance matrix for the model for visual acuity <0.1**

| | T-stage (T2) | T-stage (T3) | T-stage (T4) | Location (midperipheral) | Location (peripheral) | Optic disk V20 | Ciliary body V20 | Baseline visual acuity |
|---|---|---|---|---|---|---|---|---|
| **T-stage (T2)** | 0.270488 | 0.220763 | 0.224474 | -0.016158 | -0.007671 | -0.000005 | -0.000535 | 0.005040 |
| **T-stage (T3)** | 0.220763 | 0.270693 | 0.259249 | -0.032772 | -0.021868 | 0.000147 | -0.000976 | -0.005948 |
| **T-stage (T4)** | 0.224474 | 0.259249 | 0.336899 | -0.045208 | -0.023138 | 0.000432 | -0.000922 | 0.016381 |
| **Location (midperipheral)** | -0.016158 | -0.032772 | -0.045208 | 0.079166 | 0.066018 | 0.000296 | -0.000005 | -0.015576 |
| **Location (peripheral)** | -0.007671 | -0.021868 | -0.023138 | 0.066018 | 0.296262 | 0.000631 | -0.000462 | 0.004348 |
| **Optic disk V20** | -0.000005 | 0.000147 | 0.000432 | 0.000296 | 0.000631 | 0.000010 | 0.000000 | 0.000055 |
| **Ciliary body V20** | -0.000535 | -0.000976 | -0.000922 | -0.000005 | -0.000462 | 0.000000 | 0.000052 | -0.000204 |
| **Baseline visual acuity** | 0.005040 | -0.005948 | 0.016381 | -0.015576 | 0.004348 | 0.000055 | -0.000204 | 0.122286 |

**Supplementary table 6B: covariance matrix for the model for visual acuity <0.3**

| | Macula V20 | Central or juxtapapillary tumour location | Ciliary body V20 | Baseline visual acuity |
|---|---|---|---|---|
| **Macula V20** | 0.000006 | -0.000170 | 0.000003 | 0.000056 |
| **Central or juxtapapillary tumour location** | -0.000170 | 0.078908 | 0.000269 | -0.009459 |
| **Ciliary body V20** | 0.000003 | 0.000269 | 0.000043 | 0.000018 |
| **Baseline visual acuity** | 0.000056 | -0.009459 | 0.000018 | 0.084881 |

**Supplementary table 7A: baseline hazards for the model for visual acuity <0.1**

| hazard | time (years) |
|---|---|
| 0.0074 | 0.0192 |
| 0.0149 | 0.0219 |
| 0.0301 | 0.0383 |
| 0.0691 | 0.0411 |
| 0.0948 | 0.0575 |
| 0.1649 | 0.0602 |
| 0.1871 | 0.0767 |
| 0.1988 | 0.0794 |
| 0.2108 | 0.0958 |
| 0.2349 | 0.0986 |
| 0.2473 | 0.1068 |
| 0.2596 | 0.1095 |
| 0.2722 | 0.1150 |
| 0.2850 | 0.1342 |
| 0.2979 | 0.1369 |
| 0.3108 | 0.1752 |
| 0.3108 | 0.2190 |
| 0.3371 | 0.2327 |
| 0.3371 | 0.2519 |
| 0.3508 | 0.2875 |
| 0.3646 | 0.2902 |
| 0.3786 | 0.3450 |
| 0.3927 | 0.3751 |
| 0.4078 | 0.4052 |
| 0.4229 | 0.4216 |
| 0.4382 | 0.4435 |
| 0.4537 | 0.4983 |
| 0.4537 | 0.5010 |
| 0.4537 | 0.5366 |
| 0.4537 | 0.5394 |
| 0.4537 | 0.5722 |
| 0.4537 | 0.5749 |
| 0.4706 | 0.6270 |
| 0.4877 | 0.6571 |
| 0.5053 | 0.6680 |
| 0.5230 | 0.6708 |
| 0.5586 | 0.6927 |
| 0.5767 | 0.7118 |
| 0.5952 | 0.7310 |
| 0.6138 | 0.7337 |
| 0.6325 | 0.7502 |
| 0.6512 | 0.7556 |
| 0.6707 | 0.7830 |
| 0.6909 | 0.8350 |
| 0.7115 | 0.9199 |
| 0.7115 | 0.9747 |
| 0.7331 | 0.9829 |

| | |
|---|---|
| 0.7549 | 1.0075 |
| 0.7770 | 1.0322 |
| 0.7992 | 1.0486 |
| 0.8219 | 1.0951 |
| 0.8449 | 1.1088 |
| 0.8686 | 1.1472 |
| 0.8932 | 1.1910 |
| 0.8932 | 1.2402 |
| 0.9198 | 1.3525 |
| 0.9472 | 1.4565 |
| 0.9748 | 1.4702 |
| 0.9748 | 1.4839 |
| 1.0034 | 1.5359 |
| 1.0340 | 1.5770 |
| 1.0340 | 1.5989 |
| 1.0340 | 1.6372 |
| 1.0657 | 1.6701 |
| 1.0657 | 1.7084 |
| 1.0657 | 1.7440 |
| 1.0657 | 1.7659 |
| 1.0980 | 1.7796 |
| 1.1308 | 1.7851 |
| 1.1308 | 1.8398 |
| 1.1308 | 1.8508 |
| 1.1638 | 1.8809 |
| 1.1977 | 1.9795 |
| 1.2319 | 1.9959 |
| 1.2319 | 2.0014 |
| 1.2680 | 2.0151 |
| 1.3051 | 2.0507 |
| 1.3051 | 2.0589 |
| 1.3051 | 2.0780 |
| 1.3443 | 2.0862 |
| 1.3443 | 2.1109 |
| 1.3870 | 2.1218 |
| 1.3870 | 2.1355 |
| 1.3870 | 2.2067 |
| 1.3870 | 2.2806 |
| 1.3870 | 2.3381 |
| 1.3870 | 2.4750 |
| 1.3870 | 2.4914 |
| 1.4352 | 2.5106 |
| 1.4352 | 2.5133 |
| 1.4352 | 2.5818 |
| 1.4352 | 2.6092 |
| 1.4908 | 2.6585 |
| 1.5486 | 2.6667 |
| 1.5486 | 2.6831 |
| 1.5486 | 2.7050 |

| | |
|---|---|
| 1.5486 | 2.7242 |
| 1.5486 | 2.7406 |
| 1.5486 | 2.7625 |
| 1.6184 | 2.7981 |
| 1.6184 | 2.8556 |
| 1.6902 | 2.8693 |
| 1.6902 | 2.9541 |
| 1.7667 | 2.9706 |
| 1.7667 | 3.0000 |

**Supplementary table 7B: baseline hazards for the model for visual acuity <0.3**

| hazard | time (years) |
|---|---|
| 0.0030 | 0.0192 |
| 0.0045 | 0.0219 |
| 0.0091 | 0.0383 |
| 0.0203 | 0.0411 |
| 0.0222 | 0.0438 |
| 0.0296 | 0.0575 |
| 0.0505 | 0.0602 |
| 0.0555 | 0.0739 |
| 0.0655 | 0.0767 |
| 0.0738 | 0.0794 |
| 0.0767 | 0.0958 |
| 0.0888 | 0.0986 |
| 0.0921 | 0.1095 |
| 0.0955 | 0.1150 |
| 0.0989 | 0.1342 |
| 0.1057 | 0.1369 |
| 0.1127 | 0.1752 |
| 0.1162 | 0.1999 |
| 0.1197 | 0.2108 |
| 0.1197 | 0.2190 |
| 0.1234 | 0.2327 |
| 0.1273 | 0.2519 |
| 0.1314 | 0.4079 |
| 0.1355 | 0.4244 |
| 0.1439 | 0.4627 |
| 0.1439 | 0.5010 |
| 0.1483 | 0.5366 |
| 0.1483 | 0.5394 |
| 0.1528 | 0.5722 |
| 0.1574 | 0.5749 |
| 0.1622 | 0.5914 |
| 0.1670 | 0.6270 |
| 0.1720 | 0.6571 |
| 0.1771 | 0.6680 |
| 0.1822 | 0.6708 |
| 0.1874 | 0.6927 |

| | |
|---|---|
| 0.1927 | 0.7310 |
| 0.1981 | 0.7337 |
| 0.2035 | 0.7502 |
| 0.2090 | 0.7556 |
| 0.2147 | 0.7830 |
| 0.2208 | 0.8652 |
| 0.2271 | 0.8789 |
| 0.2334 | 0.8843 |
| 0.2334 | 0.9747 |
| 0.2398 | 0.9829 |
| 0.2466 | 0.9993 |
| 0.2537 | 1.0075 |
| 0.2608 | 1.0486 |
| 0.2681 | 1.0513 |
| 0.2755 | 1.0568 |
| 0.2833 | 1.0678 |
| 0.2913 | 1.1088 |
| 0.2997 | 1.1307 |
| 0.3087 | 1.1472 |
| 0.3182 | 1.1608 |
| 0.3280 | 1.1910 |
| 0.3280 | 1.2402 |
| 0.3383 | 1.3470 |
| 0.3486 | 1.4565 |
| 0.3486 | 1.4839 |
| 0.3594 | 1.5441 |
| 0.3704 | 1.5688 |
| 0.3704 | 1.5989 |
| 0.3704 | 1.6372 |
| 0.3704 | 1.7084 |
| 0.3704 | 1.7440 |
| 0.3704 | 1.7659 |
| 0.3828 | 1.7796 |
| 0.3955 | 1.8234 |
| 0.3955 | 1.8398 |
| 0.3955 | 1.8508 |
| 0.4089 | 1.8617 |
| 0.4228 | 1.9192 |
| 0.4371 | 1.9740 |
| 0.4518 | 1.9795 |
| 0.4518 | 2.0014 |
| 0.4678 | 2.0151 |
| 0.4840 | 2.0507 |
| 0.4840 | 2.0589 |
| 0.4840 | 2.0780 |
| 0.4840 | 2.1355 |
| 0.5022 | 2.1875 |
| 0.5022 | 2.2067 |
| 0.5022 | 2.2806 |

| | |
|---|---|
| 0.5022 | 2.3381 |
| 0.5022 | 2.4750 |
| 0.5022 | 2.4914 |
| 0.5022 | 2.5133 |
| 0.5225 | 2.5708 |
| 0.5225 | 2.5818 |
| 0.5225 | 2.6092 |
| 0.5225 | 2.6831 |
| 0.5225 | 2.7625 |
| 0.5462 | 2.7981 |
| 0.5462 | 2.8556 |
| 0.5714 | 2.8693 |
| 0.5714 | 2.9541 |
| 0.5980 | 2.9706 |
| 0.6254 | 2.9733 |
| 0.6254 | 3.0000 |

**Supplementary figure 1: Calibration plots for the final prediction models on the validation cohort**

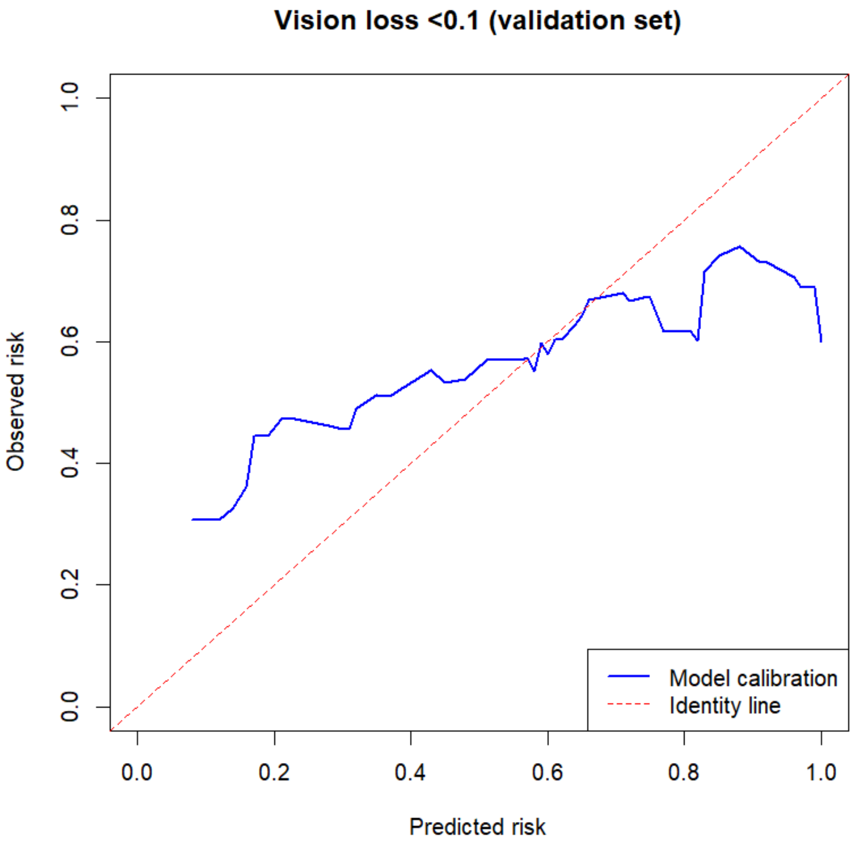


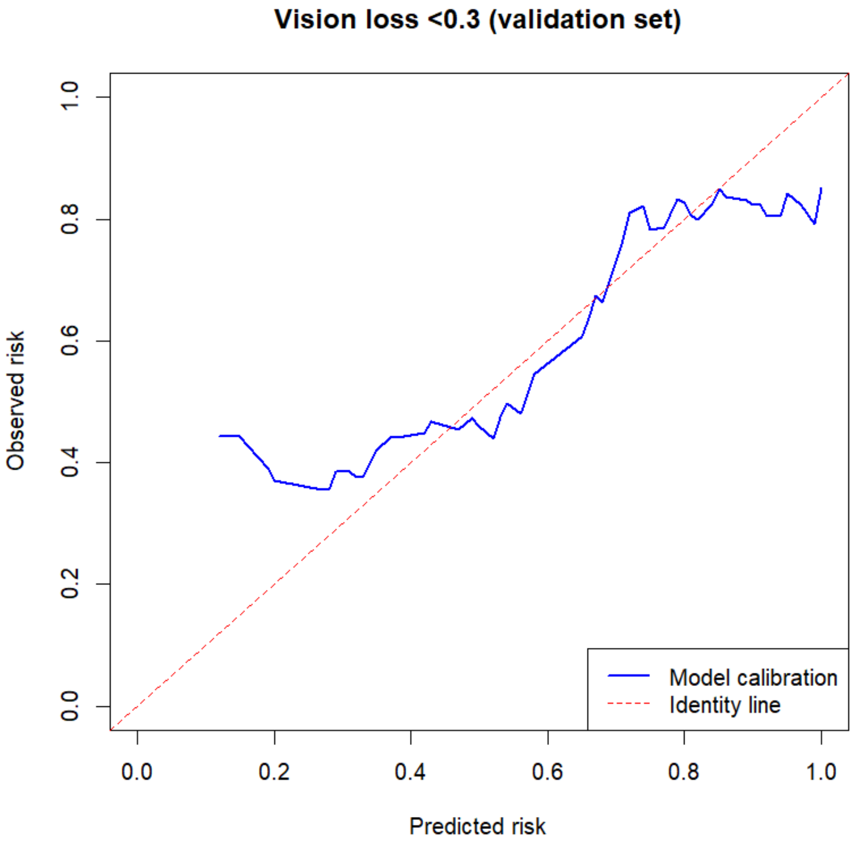
